# Implementation of Unassisted and Community-Based HIV Self-Testing (HIVST) during the COVID-19 pandemic among Men-who-have-sex-with-Men (MSM) and Transgender Women (TGW): A Demonstration Study in Metro Manila, Philippines

**DOI:** 10.1101/2021.11.13.21266310

**Authors:** John Danvic T. Rosadiño, Ronivin G. Pagtakhan, Matthew T. Briñes, Jeanno Lorenz G. Dinglasan, Denis P. Cruz, John Oliver L. Corciega, Aeronne B. Pagtakhan, Zypher Jude G. Regencia, Emmanuel S. Baja

## Abstract

**Objective:** The study aimed to demonstrate the feasibility of an unassisted and community-based HIV self-testing (HIVST) distribution model and to evaluate its acceptability among men-having-sex-with-men (MSM) and transgender women (TGW).

**Methods:** Our observational study focused on implementing the HIVST service in Metro Manila, Philippines. Convenience sampling was done with the following inclusion criteria: MSM or TGW, at least 18 years old, and had no previous HIV diagnosis. Individuals taking HIV Pre-exposure Prophylaxis (PrEP), on Antiretroviral Therapy (ART), or female sex at birth were excluded. The implementation of the study was online using a virtual assistant and delivery system via courier due to COVID-19-related lockdowns. Feasibility was measured by the number of HIVST kits successfully delivered and utilized and the HIV point prevalence rate. Moreover, acceptability was evaluated by a 10-item system usability scale (SUS). HIV prevalence was estimated with linkage to care prioritized for reactive participants.

**Results:** Out of 1,690 kits distributed, only 953 (56.4%) participants reported their results. Overall HIV point prevalence was 9.8%, with 56 (60.2%) reactive participants linked to further testing. Furthermore, 27.4% of respondents self-reported, and 13.4% of the reactive participants were first-time testers. The HIVST service had an overall mean ± standard deviation SUS score of 81.0 ± 13.0, rendering the HIVST kits very acceptable.

**Conclusions:** HIVST is acceptable and feasible to MSM and TGW. Online platforms are an innovative and effective way to deliver HIVST service during a pandemic. However, messaging to entice people to use the kit must be differentiated based on their age, gender identity and expression, and previous HIVST experience to offer the service efficiently to the target populations.

## INTRODUCTION

In 2018, the World Health Organization (WHO) recommended the use of HIV self-testing (HIVST) as an innovative approach to increase HIV testing capacity and to reach high-risk populations particularly, men-who-have-sex-with-men (MSM) and transgender women (TGW) [1]. HIVST is a WHO-recommended differentiated HIV testing option available to reach the people with HIV who do not know their status and people who have previously been diagnosed with HIV but are not on antiretroviral therapy (ART) [2]. Assisted or unassisted HIVST is done through a rapid diagnostic HIV test, using either blood or oral. It is an accurate and convenient test that respects privacy and confidentiality, outweighing current barriers to facility-based testing among key populations [3, 4].

In the Philippines, an increasing trend is observed in terms of new infections, and only about 68% of people-living-with-HIV (PLHIV) knew their status last 2020 [5], a figure far from the 90-90-90 targets of the Joint United Nations Programme on HIV/AIDS (UNAIDS) [6]. An average of 29 new cases per day was recorded in January 2021. Among the newly reported cases, 89% were MSM, with 41% from the National Capital Region (NCR) [7]. Only 28% of MSM are estimated to have undergone HIV testing and are aware of their status. With condom use during the last anal sex low among this population at 40%, they are at a higher risk for HIV compared to other key populations [8]. As a low prevalence [9-11], high incidence [6] country in HIV cases based on the estimates, reaching PLHIV who have unknown status has been difficult.

With the enactment of the Philippine Republic Act 11166, any person who engages in high-risk behavior below 15 years old is eligible for HIV testing with the assistance of social or healthcare workers, removing legislative barriers to testing among the young key population [12]. In addition, increasing HIV testing uptake among key populations is essential [13] since incidence cases are concentrated in these populations. The inadequate knowledge about HIV and the high stigma towards PLHIV among key populations also play a significant role in preventing access to testing and treatment [14]. Because of the current situation of the HIV epidemic in the Philippines, our study involved a multisectoral effort to implement an HIVST demonstration study among MSM and TGW in Metro Manila, Philippines. We aimed to demonstrate the feasibility of an unassisted and community-based HIVST model in terms of HIVST uptake, reporting rate, point prevalence, linkage to confirmatory testing by distributing HIVST kits to MSM and TGW. Furthermore, we also aimed to evaluate the usability of the kits to determine their acceptability.

## MATERIALS AND METHODS

### Study Design and Setting

This demonstration study utilized an observational study design, mainly focused on planning and implementing an unassisted and community-based model of HIVST service in Metro Manila, Philippines. Due to limitations and lockdowns caused by the coronavirus COVID-19 pandemic, the HIVST service delivery utilized the existing online channels of The LoveYourself, Inc. (TLY). This community-based, volunteer-run organization offers HIV and other sexually-health-related services in the Philippines. Moreover, an automated online messaging system was designed to gather information from target participants and deliver the HIVST kits to those identified to be eligible. The online followed a five-part flow process: expression of interest, pre-qualification process, confirmation & delivery, guided HIV self-testing process, and evaluation & post-testing process (see Figure 1 for details).

**Figure 1.**
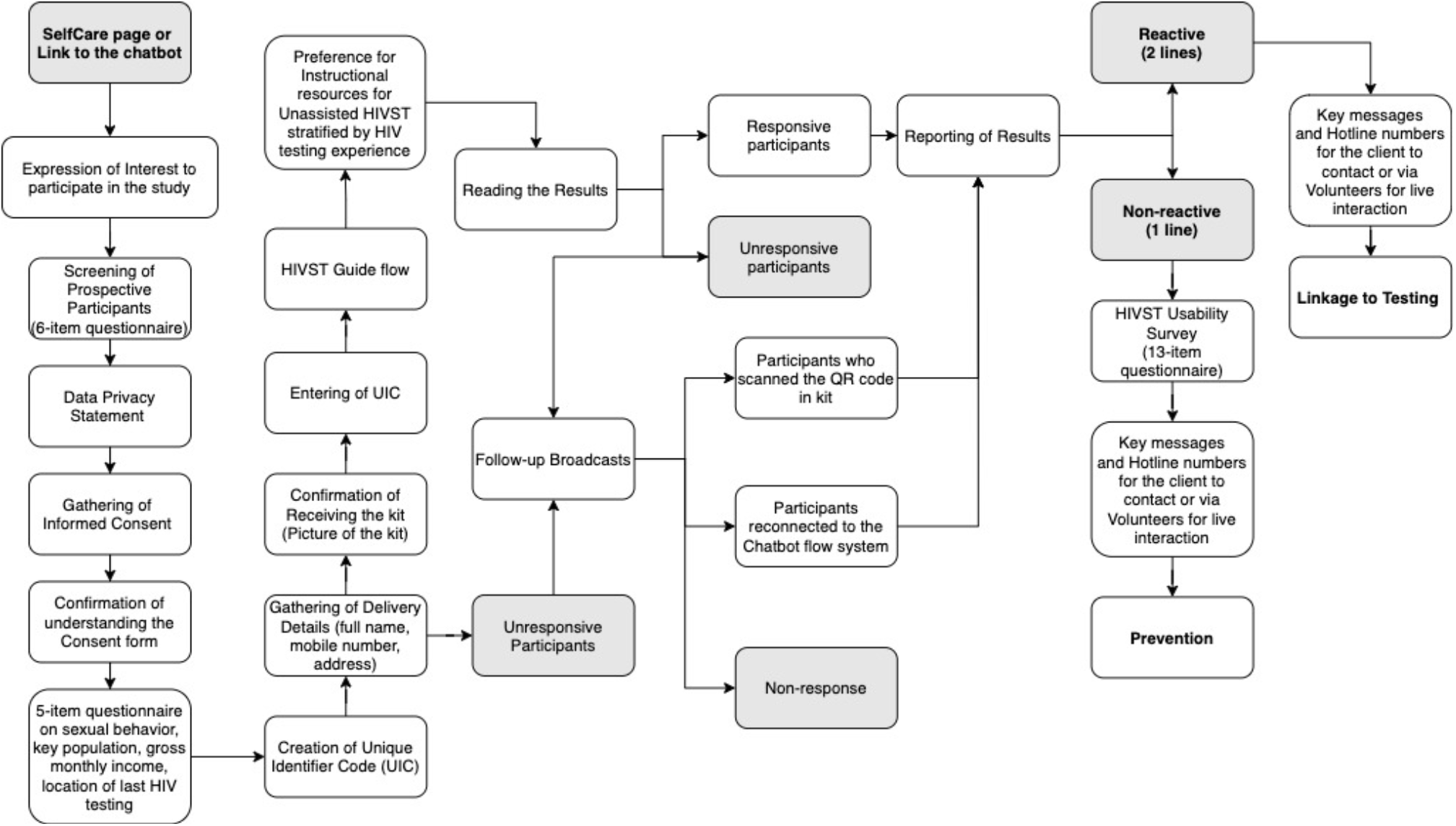
Virtual assistant flow for unassisted HIVST.

The HIVST kit (SURE CHECK® HIV 1/2 Assay USA - Chembio Diagnostics Hauppauge, New York) packaging was covered with white paper, and the kits were delivered through a private courier to maintain privacy. Furthermore, the study team monitored and validated the number of participants who accessed the virtual assistant system. The entire demonstration study was implemented from April 2020 to May 2020. Supplemental File 1 provides the instructions and guides the participants on using the HIVST kits.

### Study Participants

During the study period, individuals in the age bracket of 18 to 49 identified themselves as MSM or TGW, had no previous diagnosis of HIV, and residing in Metro Manila were included in the study. On the other hand, individuals taking Pre-exposure Prophylaxis for HIV (PrEP), on ART, and/or female sex at birth were excluded. Convenience sampling via a first-come, first-served basis was used in selecting the participants of the study.

### Data Collection and Outcome Measurements

The study utilized location-targeted Facebook advertisements to promote the study and to invite potential target participants. In addition, the study partnered with an organization that has already been offering HIV testing, prevention, and treatment services to key populations in the country. Responses were gathered through the virtual assistant and were encrypted and automatically transported to a database in real-time. At the end of data collection, the database was anonymized by assigning a unique identifier code for each client. Informed consent forms signed online by the participants were collected from the start of the study.

Socio-demographic characteristics of the participants were collected through the virtual assistant. The feasibility of delivering HIVST kits to the target key populations in a community setting was assessed based on the following parameters: the number of HIVST kits distributed (HIVST uptake), the HIV reactivity rate, and the linkage to further testing. In addition, information on the participant’s preference on the type of sources of HIVST information was also surveyed.

Furthermore, the acceptability of the HIVST kit was measured using a 10-item System Usability Scale (SUS), a standard tool that allows users to assess the usability of a given product or service [15-17]. Briefly, it is comprised of ten validated statements, which cover five positive aspects and five negative aspects of a particular tool, system, or kit. The final score is out of 100, wherein each respondent answered every question on a Likert scale from Strongly Disagree to Strongly Agree. Moreover, a SUS score of less than 50 was considered “Not Acceptable” and would imply that the HIVST kit will have usability issues. SUS scores between 50 to 70 were classified as “Marginal,” whereas a SUS score of greater than 70 was categorized as “Acceptable,” with varying degrees of usability [16]. The SUS tool was only offered to non-reactive clients through the virtual assistant.

Non-reactive participants were counseled about evidence-based preventive strategies such as the use of PrEP to decrease their risk of acquiring HIV and other sexually transmitted infections. In addition, the research team contacted participants who received invalid results and were offered to visit the nearest HIV facility for testing. For reactive clients, options were presented to access HIV testing and treatment services, and they were excluded from answering the survey.

### Data Analysis

Descriptive statistics were calculated to describe the participant’s characteristics using means and standard deviations for continuous variables and frequencies and percentages for categorical variables. SUS calculation was carried out using the formula below:

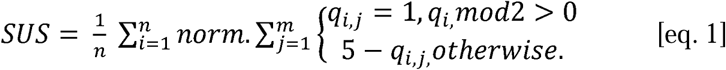

where n = number of subjects, m = 10 (number of questions), q_i,j_ = individual score per question j per subject i, and norm = 2.5 [18].

Chi-square tests were used to determine relationships between the preference of various instructional resources for HIVST and the HIV testing experience of the participants. In addition, analysis of variance was used to assess if the SUS score means stratified by age, gender identity, and HIV testing experience were significantly different among the stratified characteristics. All statistical analyses were carried out using STATA 17 (www.stata.com) software. A p-value of ≤ 0.05 was considered statistically significant.

### Ethical considerations

The University of the Philippines Manila Research Ethics Board (UPMREB) has approved the ethical clearance of this study with UPMREB CODE: 2019-474-01. All data were treated following the Philippine Data Privacy Act of 2012 (Republic Act 10173).

## RESULTS

### HIVST Cascade

The cascade of the feasibility of delivering HIVST kits to the target participants is presented in Fig. 2. These numbers were combined data from the virtual assistant analytics generated at the end of the study period. Out of 4,205 individuals who submitted a message, 4,163 (99%) individuals expressed their interest in getting the HIVST kit, but only 2,543 (60%) were eligible to receive the kit. A total of 1,690 HIVST kits were distributed in this study, but only 953 participants reported their results (reporting rate = 56.4%). The observed HIV point prevalence from April 2020 to May 2020 using the HIVST kits was 9.8% (93/953). Among the reactive participants, 60.2% (56/93) were linked to further testing. Non-reactive participants were asked to answer the SUS survey, and 93.9% (797/849) sent their responses. Characteristics of the respondents who received the kits, who reported their HIV results, and who answered the survey are presented in Table 1.

**Figure 2.**
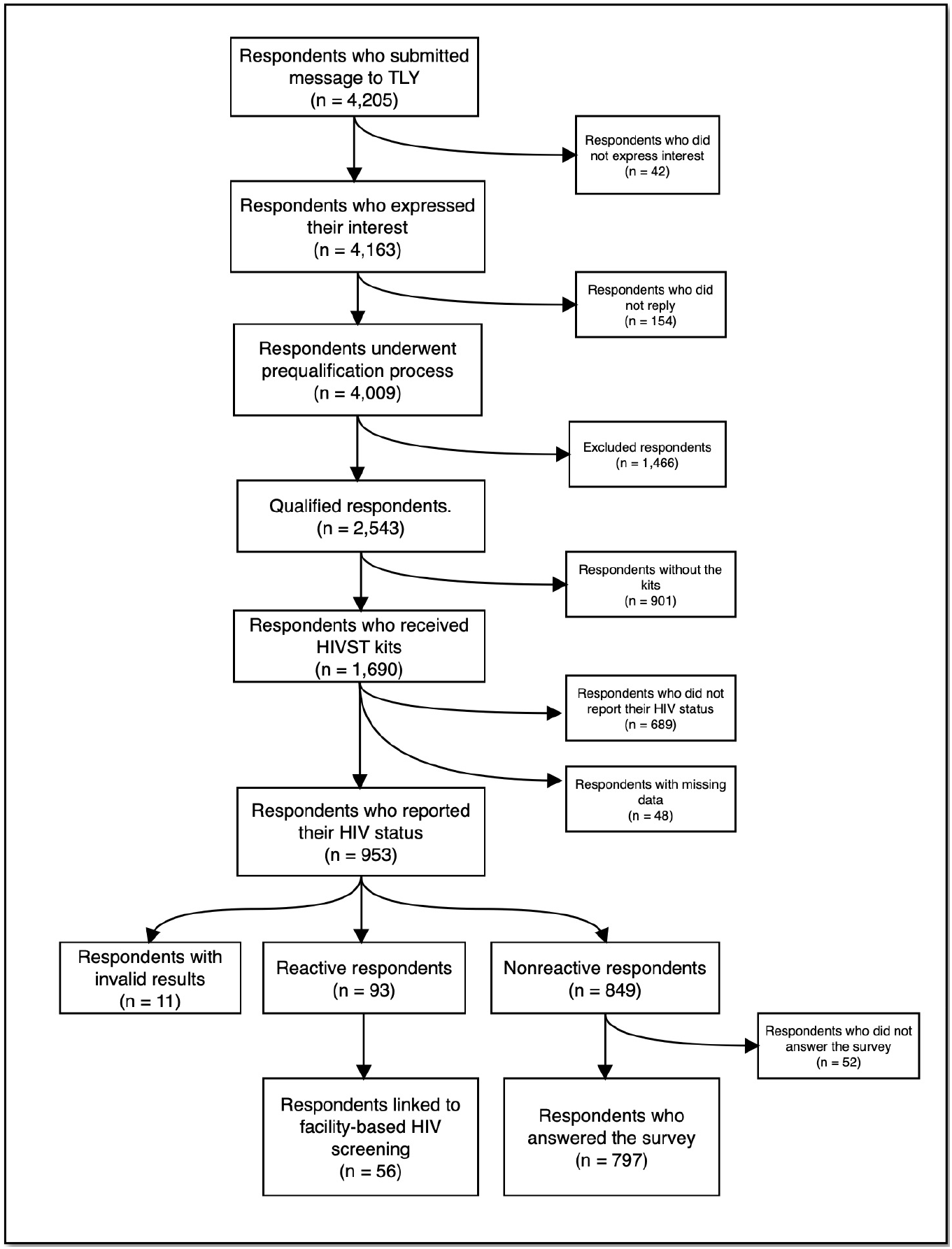
HIVST feasibility cascade chart.

**Table 1.** Characteristics of the study population.

| **Characteristics <sup>a</sup>** | **Received the HIVST kit**<br>**(n = 1,690)** | **Reported the results of the HIV test**<br>**(n = 953)** | **Answered the online survey**<br>**(n = 797)** |
| --- | --- | --- | --- |
| Age [mean (SD)] | 28.1 (5.3) | 27.9 (5.2) | 28.0 (5.3) |
| 18 to 24 years old | 452 (26.7) | 277 (29.1) | 231 (29.0) |
| 25 to 35 years old | 984 (58.2) | 559 (58.6) | 459 (57.6) |
| 36 years old & above | 204 (12.1) | 115 (12.1) | 105 (13.2) |
| Missing data | 50 (3.0) | 2 (0.2) | 2 (0.2) |
| Gender identity |  |  |  |
| MSM | 1,531 (90.6) | 892 (93.6) | 744 (93.4) |
| TGW | 38 (2.2) | 19 (2.0) | 16 (2.0) |
| Prefer not to say | 70 (4.2) | 40 (4.2) | 35 (4.4) |
| Missing data | 51 (3.0) | 2 (0.2) | 2 (0.2) |
| Previous testing experience |  |  |  |
| Got tested | 1,187 (70.2) | 692 (72.6) | 585 (73.4) |
| Never tested | 454 (26.9) | 261 (27.4) | 212 (26.6) |
| Missing data | 49 (2.9) | - | - |
| Testing site commonly used |  |  |  |
| TLY | 731 (43.3) | 426 (44.7) | 372 (46.7) |
| Social Hygiene Clinics | 144 (8.5) | 79 (8.3) | 68 (8.5) |
| Private Clinic | 91 (5.4) | 50 (5.2) | 43 (5.4) |
| Hospital (Private/Public) | 114 (6.7) | 65 (6.8) | 48 (6.0) |
| CBS Motivator | 25 (1.5) | 17 (1.8) | 16 (2.0) |
| Other centers | 82 (4.9) | 55 (5.8) | 38 (4.8) |
| Never tested | 454 (26.9) | 261 (27.4) | 212 (26.6) |
| Missing data | 49 (2.9) | - | - |
| Monthly Income, PHP |  |  |  |
| < 10,000 | 144 (8.5) | 75 (7.9) | 53 (6.7) |
| 10,000-19,999 | 377 (22.3) | 198 (20.8) | 162 (20.3) |
| 20,000-29,999 | 338 (20.0) | 202 (21.2) | 173 (21.7) |
| 30,000-49,999 | 236 (14.0) | 142 (14.9) | 127 (15.9) |
| 50,000-99,999 | 99 (5.9) | 58 (6.1) | 49 (6.2) |
| 100,000 and up | 43 (2.5) | 27 (2.8) | 23 (2.9) |
| Not applicable<sup>b</sup> | 175 (10.4) | 114 (11.9) | 99 (12.4) |
| Prefer not to say | 229 (13.6) | 137 (14.4) | 111 (13.9) |
| Missing data | 49 (2.9) | - | - |
| HIV test result |  |  |  |
| Reactive | - | 93 (9.8) | - |
| Non-reactive | - | 849 (89.1) | 797 (100.0) |
| Invalid | - | 11 (1.1) | - |
| SUS score [mean (SD)] | - | - | 81.0 (13.0) |

<sup>a</sup> Distributions of variables are reported as n (%) unless specified otherwise; <sup>b</sup> Includes students and unemployed.

In Table 1, the mean age of the respondents who received the kits, who reported their HIV results, and who answered the survey was similar (∼28 years old). Most of them were MSM (∼93%) across all groups, and only about ∼2% were TGW. Moreover, 1 out of 5 respondents had an average monthly income between PhP 10,000-19,999, while almost 3% of them earned > PhP 100,000 monthly. In addition, the distribution of the respondent’s characteristics who received the kits, who reported their HIV results, and who answered the survey did not change at each stage of the HIVST cascade. This similar distribution implies that the respondents who answered the survey may be considered representative of the study’s target population. Furthermore, the overall SUS score was 81.0, which renders the kits very usable and acceptable.

Of the 56 reactive participants linked to further testing, approximately 2 out of 3 belonged to the 25 to 34 years old age group (66.1%), and 19 out of 20 were MSM (94.6%). Moreover, among the 261 first-time testers, the estimated HIV point prevalence was 13.4% (35/261) (see Table 2 for details).

**Table 2.** Characteristics of reactive respondents linked to further testing (n=56).

| Characteristics <sup>a</sup> | Values |
| --- | --- |
| Age groups (years) |  |
| 15 to 24 | 17 (30.3) |
| 25 to 34 | 37 (66.1) |
| 35 to 49 | 2 (3.6) |
| Gender Identity |  |
| MSM | 53 (94.6) |
| TGW | 0 (0.0) |
| Preferred not to say | 3 (5.4) |
| Testing Experience |  |
| Never Tested | 35 (62.5) |
| Ever Tested | 21 (37.5) |

<sup>a</sup>Distributions of variables are reported as n (%).

The respondents’ preferences when it comes to sources of instructional materials on HIVST were also collected and summarized. More than half of the participants preferred to use instructional videos (51.9%) and printed materials or inserts (52.4%). This preference is true regardless of the HIV testing experience of the respondents. In contrast, a landline hotline option was the least preferred among all types of resources (3.2%). In addition, no difference in the preference of various sources of instructional materials was observed between ever-tested and never-tested participants (p-values > 0.05) (see Table 3 for details).

**Table 3.** Preference of instructional resources stratified by HIV testing experience (N = 792).

| Resources | Total<br>n = 792 |  | Ever Tested<br>n = 584 |  | Never Tested<br>n = 208 |  | *p*-values |
| --- | --- | --- | --- | --- | --- | --- | --- |
|  | n | % | n | % | n | % |  |
| **Instructional videos (%)** |  |  |  |  |  |  | 0.33 |
| Yes | 411 | 51.9 | 297 | 50.9 | 114 | 54.8 |  |
| No | 381 | 48.1 | 287 | 49.1 | 94 | 45.2 |  |
| **Printed Materials/Inserts (%)** |  |  |  |  |  |  | 0.63 |
| Yes | 415 | 52.4 | 309 | 52.9 | 106 | 51.0 |  |
| No | 377 | 47.6 | 275 | 47.1 | 102 | 49.0 |  |
| **Talking with celebrity endorsers**<br>**via Messenger (%)** |  |  |  |  |  |  | 1.00 |
| Yes | 145 | 18.3 | 107 | 18.3 | 38 | 18.3 |  |
| No | 647 | 81.7 | 477 | 81.7 | 170 | 81.7 |  |
| **Hotline (%)** |  |  |  |  |  |  | 0.11 |
| Yes | 25 | 3.2 | 15 | 2.6 | 10 | 4.8 |  |
| No | 767 | 96.8 | 569 | 97.4 | 198 | 95.2 |  |

### HIVST Kit Usability Score

The HIVST kit used in the demonstration study had an overall usability score [± standard deviation (SD)] of 81.0 (± 13.0). SUS scores were further stratified based on age groups, gender identity, and HIV testing experiences. The SUS score means differ significantly among the three age groups (p-value < 0.01), with the 35 to 49 age group having the highest SUS score mean (83.6 ±11.6) and the 15 to 24 age group having the lowest SUS score mean (78.1 ±13.1). For gender identity, MSM participants had higher SUS scores (81.1 ± 13.0) than TGW and other participants who preferred not to disclose their gender identity, but the means of the SUS score was not significantly different (p-value = 0.25). Lastly, participants who had prior HIV testing experience had higher SUS scores (81.5 ± 12.2) compared with never-testers (79.4 ±15.0) with the SUS score means significantly different (p-value = 0.04). All observed SUS scores were under the “acceptable” category, rendering the HIVST kit usable (see Table 4 for more details).

**Table 4.** HIVST kit SUS scores, stratified by participants’ characteristics (N = 783).

| Participants stratified by characteristics | n | Mean Usability Scores (±SD) | *p*-values |
| --- | --- | --- | --- |
| Overall | 783 | $81.0 \pm 13.0$ | |
| Age groups (years) |  |  |  |
| 15 to 24 | 225 | $78.1 \pm 13.1$ | <0.01 |
| 25 to 34 | 451 | $81.7 \pm 13.1$ | |
| 35 to 49 | 104 | $83.6 \pm 11.6$ | |
| Gender Identity |  |  |  |
| MSM | 731 | $81.1 \pm 13.0$ | 0.25 |
| TGW | 15 | $80.7 \pm 13.8$ | |
| Preferred not to say | 35 | $77.4 \pm 12.3$ | |
| HIV Testing Experience |  |  |  |
| Never Tested | 206 | $79.4 \pm 15.0$ | 0.04 |
| Ever Tested | 577 | $81.5 \pm 12.2$ | |

## DISCUSSION

Similar to other countries, HIV is still associated with stigma and discrimination in the Philippines. The Philippines was recording 29 new cases per day as of January 2021, with the MSM population among the majority of the newly diagnosed. Innovative ways of reaching this population for testing and educating, especially during a pandemic, is one strategy for controlling the large-scale generalized HIV epidemic in the country among the high-risk and at-risk groups. Confounding factors such as low condom use, unsafe injecting drug use practices, lack of sexual health education and misconception about HIV/AIDS, and religion and conservatism further increase the rate of infection in the country [19]. The fear of testing positive prevents access to HIV testing, and some would prefer to die than face judgment from other people. In a study in Singapore, HIVST’s advantage of being completely anonymous, ease of access, and autonomy, made it preferable, with an inclination towards blood-based testing. However, the participants did not feel confident in using blood-based testing due to fear of pricking themselves, which they felt could affect the result [20].

In our study, anonymity and confidentiality generated a high demand for the HIVST kit, with more than 4000 expressing interest. There was a recorded 56.4% reporting rate and a 9.8% reactivity rate among those eligible to receive the kit. The majority of those reported were in the 25-35 years old age group and with testing experience. Other studies on the feasibility and acceptability of HIVST kits were conducted in Africa [21-25] and elsewhere [26-28]. In a study done in Africa, they compared the usability of the oral and fingerstick methods of HIVST. The participants reported the kits to be easy to use, especially those with previous testing experience. Inclusion of visual instructions such as videos or photos and simple sample collection reduced errors in using the kits, especially in interpreting results. They also found the oral test easier to use but preferred the fingerstick test, which may improve with further use and instructional resources [22, 23]. Following the findings in previous studies [23, 29, 30], instructional materials were included in our HIVST package. This inclusion of instructional materials made the kit even more accessible as this facilitated learning to use the kit easier. In addition, the participants preferred to access the instructional videos found online to guide them in using the kit and interpreting the results. However, in a limited resource setting country like the Philippines, internet access could still be a challenge in some areas, which made the second preference of instruction to use was the printed materials/inserts.

The median ages for participants who took HIVST from other studies [28, 31, 32] were found to be in agreement with our study. In addition, the kit’s usability score and the user’s age was significantly related, with observed higher usability score among the 35 to 49 age group (83.6) compared with the 15 to 24 age group (78.1), which could be indicative of the comfortability and confidence of the older age group in using the kits. Our result could also be meaningful of the purpose of getting the HIVST kit. The younger age group’s purpose could primarily be exploration and curiosity regarding the self-testing kit, whereas the older age group, more critical, could mostly be health-related.

Implementing HIVST in a community-based setting [28, 31-35] in partnership with different stakeholders is a common feature across studies on HIVST conducted online [28, 31, 32] since reactive participants can be further linked to testing and treatment easily, reflecting the central goal of HIVST [1, 36]. A review article summarized the implementation outcomes for HIVST that would help implementers assess the program’s success. They found HIVST acceptable and appropriate, perceived convenient, and better at maintaining confidentiality than standard testing. However, the lack of counseling and linkage to care, the occurrence of user errors, and cost-effectivity were the usual concerns [21].

Our demonstration study maximized the usage of the virtual assistant from ordering the kit to the reporting of the results and linking the reactive participants to confirmatory testing and treatment. Video materials were included in the system for more information on HIV treatment and prevention. Moreover, the kit consists of instructions for use, including a guide on how to use the kit and contact details to ensure participants are well guided on their next steps should they get a reactive result. Uptake of HIVST kits, HIV point prevalence, number of first-time testers, and usability of the HIVST kit were the leading indicators for the success of our study implementation. Our study distributed a more significant number of kits across the target populations, albeit with higher findings on point prevalence. The first-time testers who reported their results in our study were similar to another research finding [31]. Furthermore, the primary goal of our study was to provide evidence that HIVST can capture first-time testers among members of the key populations (MSM and TGW) who have undiagnosed HIV. In addition, the results of our HIVST implementation suggest feasibility in the Philippines.

Association studies linking the different socio-demographic variables and sexual behaviors, and the HIV status the participants mainly were seen on other feasibility and acceptability researches on HIVST kits [31, 32, 37-39]. One study reported that among first-time testers who had reactive results, having multiple sexual partners and reporting inconsistent condom use were associated with an HIV-positive result [31]. In contrast, our study did not require reactive participants to answer questions on HIVST usability because linking them to further confirmatory testing was more prioritized than getting insights from the reactive participants. Therefore, further studies may be needed to look at the predictors of HIVST among reactive first-time testers in the Philippines, particularly the MSM and TGW populations, as supported in our results.

The study also included the TGW population since they are also considered a high-risk group in the Philippines for acquiring HIV. We were able to reach this target population but only captured a modest sample size. This finding is an important observation since TGWs were considered high-risk for HIV with a high global pool prevalence of HIV (17.7%) among TGW in low- and middle-income countries [40]. This high global pool prevalence of HIV is evidenced by high transmission probability of unprotected receptive anal intercourse consistently engaged by TGW, like most MSM [41, 42], in addition to consistently rare access to appropriate clinical prevention, treatment, or care services specifically in low- and middle-income countries [43, 44]. Nevertheless, our study provided data regarding uptake of HIVST among TGW. Future research must focus on the design to disseminate demand creation strategies for TGW and increase their participation in the HIVST to generate evidence relevant to their community.

In terms of acceptability, HIVST kit usage in other studies were found to be usable across the key populations, particularly on MSM and TGW [4, 38, 39, 45, 46], which is similar to the findings from our study. In addition, reports on social harm were not observed during the implementation of our study, a result comparable in other studies [47-49], while other studies reported otherwise [50-54]. However, stigma still plays a role in HIV diagnosis in the Philippines, and self-harm could still be possible after receiving a diagnosis. Partnering with organizations and other institutions that provide counseling and quick linkage to care and treatment would help prevent self-harm. In our study, the participants were given options to contact a center using the virtual assistant and the materials included in the kit.

HIVST offers a discreet, convenient, and empowering way for those who may not otherwise test in a facility [47]. The findings of our study showed that HIVST is effective in reaching those who want to know their HIV status. Since the low uptake of HIVST was observed among the young key population and first-time testers in our study, an opportunity for future implementers to capitalize on creating materials for HIVST targeted at these populations is warranted. Key messages must be different depending on the client’s age bracket and HIV testing experience. For clinicians, our study provided evidence regarding demand, uptake, and acceptability of HIVST among MSM and TGW in the Philippines that can help them be informed in deciding how to offer HIVST to their patients. For implementers, the design of our demonstration study can be patterned to future programs on HIVST that are tailored to other local MSM and TGW communities in the Philippines. HIVST is said to increase HIV testing [47, 55, 56] among key populations; thus, implementers should include HIVST as one of the key indicators in their existing HIV Testing and Counselling Services (HTS). Lastly, funding agencies or companies may consider HIVST their Corporate Social Responsibility (CSR) service by helping the community as potential sustainable funding sources [57]. Some countries in Africa scaled up HIVST distribution through catalytic donor investments [58, 59].

The delivery of HIVST kits to the MSM and TGW populations is feasible as reflected by high HIVST kit uptake, reporting rate, point prevalence, and reactive participants linked to further testing. Most reactive participants were linked to testing and treatment through TLY, while three participants were linked to other treatment hubs. TLY has a testing and treatment facility in key locations in the metro, which made the recruitment process for this study easier. Moreover, participants who tested reactive were immediately linked to testing and treatment with the help of volunteer TLY counselors, decreasing the cost of study for post-counseling services. Using a virtual assistant to deliver HIVST kits during the COVID-19 pandemic, the research team protected the privacy and confidentiality of interested respondents and participants. Various options on how to access instructional materials to use the HIVST kit were also provided (e.g., through the virtual assistants or the materials included with the kit), ensuring participants’ autonomy.

Mobility limitations due to the COVID-19 pandemic decreased the number of HIVST kits that can be delivered primarily in cities in Metro Manila that were inaccessible due to enhanced community quarantines and lockdowns. Due to the high demand for the HIVST kits, the research team faced difficulty to follow-up participants who did not report their results. In addition, participants were not required to answer all questions if they felt uncomfortable. Thus, decreasing the data available for analysis. Since the study was conducted online, participants who did not have access to the internet were not reached.

Our study, to our knowledge, is the first in the Philippines that look at the feasibility and acceptability of HIVST kits on a larger scale. Furthermore, our study findings cannot be generalizable to other high-risk and at-risk populations. Future studies are needed to explore the feasibility and acceptability of HIVST in the general population, including other key populations such as female and male sex workers, people who inject drugs, and young key populations.

Additional items on demographic questionnaires and questions on condom use and sexual behaviors can be included to characterize further those who want to use the HIVST kit. Moreover, barriers and facilitators considered in accessing HIV testing and treatment must also be ascertained among participants since these could potentially help design HIVST programs effectively and efficiently in the future. Finally, in distributing HIVST kits to clients, implementers can explore other means of distributing kits through pharmacies, social hygiene clinics, etc., to reach those who do not have internet access [60, 61].

As we increase HTS coverage, the proportion of HIV-positive diagnoses will decrease across all approaches, necessitating a more strategic and focused method to achieve consistent or higher levels of positivity for all HTS approaches, including HIVST [57]. HIVST can also contribute to a more comprehensive provider-initiated HTS in public clinics that are highly congested with a limited testing capacity or poorly managed [62, 63].

## CONCLUSION

Reaching the vulnerable population for HIV testing is still crucial in achieving the 90-90-90 target of UNAIDS beyond 2020. MSMs and TGW are the key populations that are greatly affected by the COVID-19 pandemic. Introducing new HIV testing modalities, such as HIVST, is vital to reach more people in knowing their HIV status and access treatment or prevention packages. Our demonstration study showed that implementation of HIVST is feasible, and the HIVST kits used were generally acceptable to the MSM and TGW populations. Implementers must consider the utilization and strengthening of online platforms during a pandemic because this is an effective way to reach and offer HIVST and other services to the hard-to-reach populations. In addition, implementers must also consider the messaging of all materials based on age, gender identities and expression, and HIV testing experience to entice everyone to use HIVST kits. Even if HIVST is an effective tool in reaching the population, there have been a lot of inhibitions when it comes to this innovation. The national government must create a national guideline on implementing HIVST in the country, considering the feasibility and acceptability of HIVST and those who have doubts and concerns about implementing this innovative service. This study shows that HIVST is a promising alternative in knowing a person’s HIV status especially, to control the large-scale generalized HIV epidemic in the country among high-risk groups. This study is a proof-of-concept that there are more gains for the Philippines to begin the implementation of this innovation.

## Data Availability

The data analyzed are not publicly available due to the risk of identifying individual participants.

## Acknowledgments

The research team gratefully acknowledges the contributions of many individuals in the study.

First of all, we would like to thank **Mohan Anasalam** of Chembio Asia Pacific, **Jorelyn Pinuela** of Isopharma, Inc. and **Dr. Gundo Weiler** of World Health Organization, Philippines for generously giving us the resources to accomplish the study, to **John Darwin Ruanto and Edgar Daniel Bagasol** for helping us in the administration and funding of these resources.

A million thanks to **Paul Victor Junio, Lord-Art Lomarda**, **Raybert Domingo, Ria Briñes** and **Kurt Silvano** for the creation of promotional materials and online systems to help us gather respondents and organize data.

We would also like to thank the LoveYourself Life Coaches and case management team namely **Jose Mari Maynes, Emery Mandel, Marlo Bryan Galvez, Tyrone Cudeldiego, Ronald Bugarin, Raymond Martin Manahan, Joy Daguiso, Marvin Frondoza, Roberto Pinauin, Harrold Abilas, Leo Pura, Michael Dela Paz, Aisia Jesse Castelano** and **Eda Catabas** for helping us in addressing the concerns of the participants as they learn more about HIV prevention or treatment packages.

The team would also like to express our gratitude to **Mark Ryan Costales, Antonio Latorino, Raphael Kevin Tevar, Queenie Sheena May Mauhay**, **Reinalyn Espiritu and Lady Lee Callejo** for helping us in administering the delivery of necessary materials to the participants. Also, to **Rolando Bello Jr. and LoveYourself Rye-ders** namely **Sir Galo, Reimil, Jayson, Cha, Jenevive, Marco, JR, Dhem, Joel, Fernando, Niel and Caloy** for delivering the materials with full confidentiality to our respondents and participants and to **Jose Buendia Jr**., **Richard Bello, Arvin Salayon and AcXess by LoveYourself** for taking care of our clients as they visit the facilities to access life-saving HIV treatment.

Thank you to our advocates and ambassadors, **Miss Universe 2018 Catriona Gray** and **Paolo Gumabao** for sharing their time and showing their support to HIV advocacy.

Lastly, **to our 1**,**100 LoveYourself volunteers** who helped us in spreading these innovations to the community and to **and to all our respondents** who generously shared their insights and thoughts to help us create ripples of positive change in the community.

## Contributors

RGP, JTR and ESB conceptualized and led in the direction of the whole study. MTB, ZGR, JTR and ESB contributed to the protocol development. ABP, JTR, JLC and MTB handled all the necessary legal approvals needed in order to push through with the study. JGD and MTB collected and managed the data of all participants. MTB JGD and JTR finalized all the given data before analysis. JTR, ZGR, ESB analyzed the data. ESB, MTB, JGD, ZGR and DPC wrote the first draft of the manuscript. All authors helped with the revision of the manuscript. All authors have agreed on the final version of the manuscript and are responsible for all aspects of this study, thus ensuring its accuracy and integrity.

