## Supplemental File 1 for "Implementation of Unassisted and Community-Based HIV Self-Testing (HIVST) during the COVID-19 pandemic among Men-who-have-sex-with-Men (MSM) and Transgender Women (TGW): A Demonstration Study in Metro Manila, Philippines"

**Supplemental File 1:** Instructions for Use (HIV Self-Testing Kit)

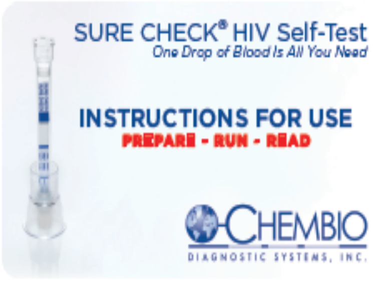


### •• READ ME NOW ••

SURE CHECK HIV Self-Test is a screening test for HIV (the virus responsible for AIDS).

SURE CHECK HIV Self-Test is intended for use at home in a private setting and therefore is not a diagnostic result. Please ensure you follow up with a healthcare professional.

For single-use only. Do not open foil pouch containing device until ready to test.

Please carefully read all of the following instructions prior to using the test.

You will need a watch, clock, or other timing device.

Wash hands prior to running test.

Run test in a well-lit area.

Use within expiration date.

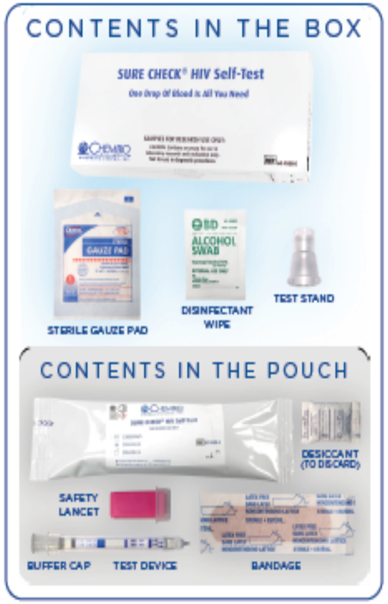


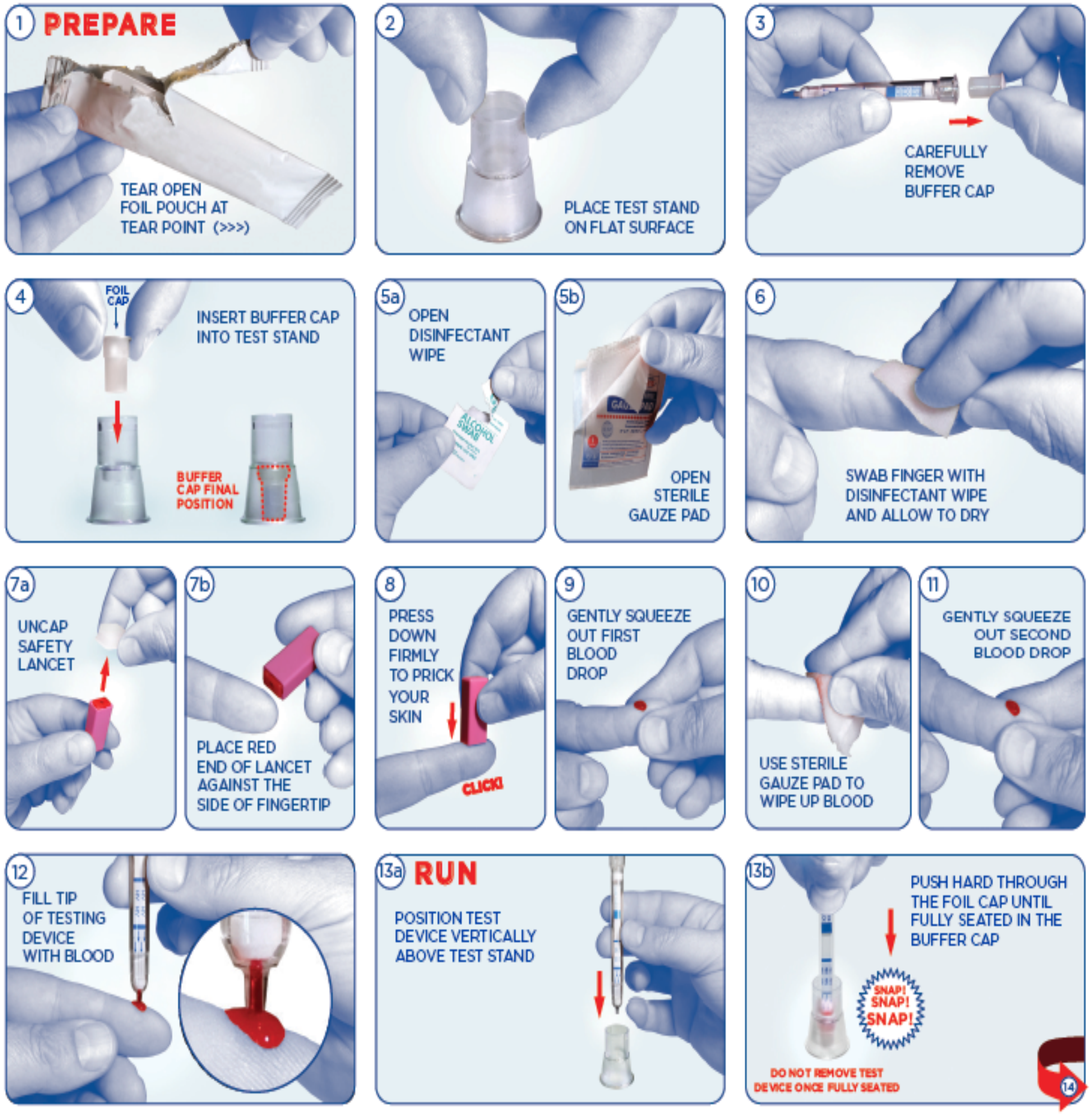

14

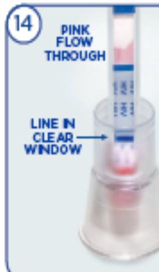


CHECK FOR PINK FLOW THROUGH WITHIN 1 MINUTE. IF NO COLOR FORMATION IS OBSERVED, PUSH DEVICE FURTHER DOWN INTO BUFFER CAP

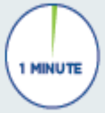


15a

WAIT 15 MINUTES FOR RESULTS

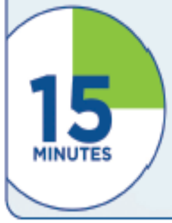


15b

READ RESULTS BETWEEN 15 AND 20 MINUTES

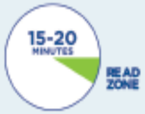


DO NOT EXCEED 20 MINUTES BEFORE READING RESULTS

16a

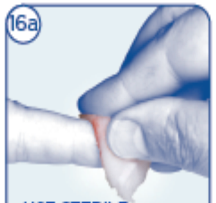

USE STERILE GAUZE PAD TO CLEAN FINGER

16b

APPLY BANDAGE TO FINGER

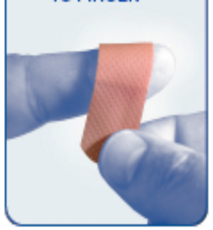

17 **READ**

COMPARE YOUR TEST DEVICE RESULTS TO THE POSSIBLE OUTCOMES

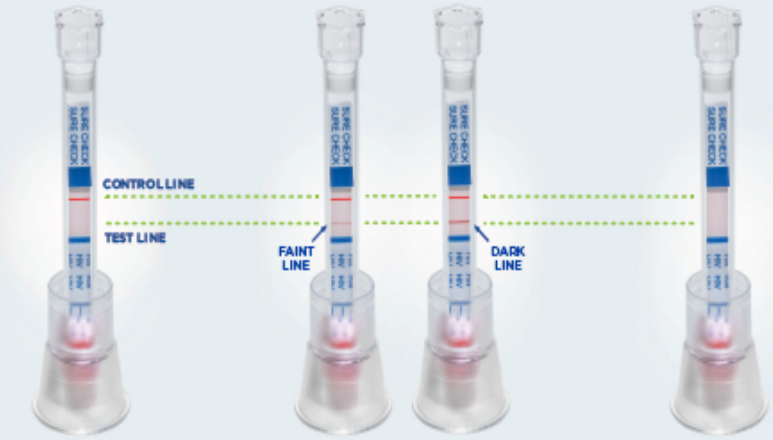


NEGATIVE POSITIVE INVALID

### NEGATIVE RESULTS

If test shows ONLY the Control Line, your test result is NEGATIVE.

**Test again in 3 months.**

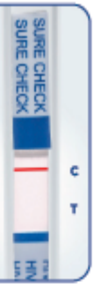


### POSITIVE RESULTS

If test shows BOTH the Control Line AND Test Line, your test result is POSITIVE.

**You MUST have a POSITIVE RESULT confirmed by a healthcare professional.**

Protect yourself and others.

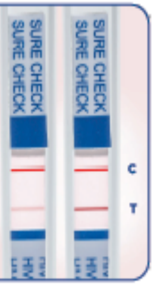


### INVALID RESULTS

If your results do not match negative or positive, the result is INVALID. It is necessary to run another test.

If repeated invalid results occur, consult a healthcare professional.

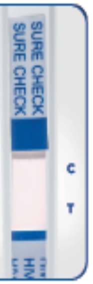


### •• LEGEND OF SYMBOLS ••

- IVD — In vitro diagnostic
- Do not reuse
- Expiration Date:
- Store at 8°-30°C
- Carefully read Instructions For Use
- Buffer Cap Warning:
  Contains sodium azide ≥ 0.2%; Harmful if ingested. Very toxic gas released upon contact with acid. Harmful to aquatic life, with long lasting adverse effects. Contains gentamicin sulfate. May produce an allergic reaction. Avoid all contact with eyes, skin and clothing. Remove spills.

Manufactured by Chembio Diagnostic Systems, Inc.
3661 Horseblock Road, Medford, NY 11763 USA
1-844-243-6246
*www.Chembio.com*
**

### •• WARNINGS AND •• PRECAUTIONS

Test must be used with fresh sample of blood taken from finger.

This test may not detect the most recent HIV infections (e.g. exposure to HIV in the 3 months prior to using the test). If you have been exposed to HIV within the past 3 months, a negative result may not be accurate.

The test kit must be stored within the specified storage temperatures of 8° to 30°C (46° to 86°F) to ensure proper performance of the test.

This test is not suitable if you are receiving antiretroviral treatment for HIV.

If your result is positive you should contact your local sexual health clinic or healthcare professional who will perform a confirmatory HIV test and provide advice.

If your test is negative it does not mean you are definitely not infected with HIV, especially if your exposure may have occurred within the past 3 months. If you engage in activities that increase your risk of exposure to HIV you should test regularly.

To dispose of your self-test please place all of the components back into the box. The box should be shut to help protect your privacy and thrown away with your normal rubbish.

Used safety lancets might be classified as medical waste by health authorities in your area. To reduce the risk of injury from a used lancet, please follow local requirements for its disposal.

For further information on HIV Self-Testing visit *www.HIVST.org*

SURE CHECK® is a registered trademark of Chembio Diagnostic Systems, Inc.

©2018 Chembio Diagnostic Systems, Inc., all rights reserved.

10-1422-0 Rev 2
Product Code: 60-9508-0
Effective Date: May, 2018
